# Quantitative Facial Phenotyping and Age-Associated Differences in Sickle Cell Disease Using Interpretable Machine Learning

**DOI:** 10.64898/2026.03.03.26346563

**Authors:** D’Andre Spencer, Xinyang Liu, Kizito Mosema-Be-Amoti, Gisèle Kandosi, Matthew S Bramble, Faisal Al Munajjed, Esther Likuba, Daniel Okitundu-Luwa E-Andjafono, Laure Tshibambe, Brian Colwell, Kristen Howell, Nicole O’Brien, Christopher Moxon, Syed Muhammad Anwar, Antonio R Porras, Dieudonne Mumba Ngoyi, Eric Vilain, Desire Tshala-Katumbay, Marius George Linguraru

## Abstract

**Background:** Sickle cell disease (SCD) is a common inherited genetic disorder and contributor to global childhood mortality and morbidity, particularly in Sub-Saharan Africa. Craniofacial and skeletal changes have been described in SCD, but the subtle soft-tissue facial patterns and their variation have not been quantitatively characterized using machine-learning methods.

**Methods:** We analyzed 308 front-facing smartphone photographs from 154 participants with electrophoresis-confirmed SCD and 154 age- and sex-matched controls aged from 5 months to 19 years recruited in Kinshasa, Democratic Republic of the Congo. Predefined geometric and texture-based facial descriptors were extracted from standardized images using manually annotated landmarks. An interpretable linear support vector machine with recursive feature elimination was evaluated using leave-one-out cross-validation, with features ranked according to their relevance for distinguishing participants with SCD from controls. Cross-sectional age- and sex-stratified analyses evaluated variation in selected features

**Results:** The facial analysis algorithm identified eight geometric and six texture discriminative features that were significantly different between the cohorts. A combination of six geometric features contributed most consistently to internal discrimination: distance between medial and lateral canthi, angle at nasal ala, distance from nasion to philtrum, distance from medial canthi to the columella, distance from columella to the lower lip, and distance between nasal alae. The model achieved an AUROC of 0.843 [95% CI: 0.798,0.887], and an accuracy of 0.795 [95% CI: 0.750,0.840]. Between-group difference was limited in the youngest age groups and were more pronounced in several older age groups. **Conclusion:** These findings demonstrate preliminary evidence that SCD is associated with quantifiable morphometric patterns. External, longitudinal, and clinically characterized studies are needed to determine whether these features may serve as reproducible digital biomarkers within a broader supportive disease-assessment framework.

**Author Summary:** Sickle cell disease (SCD) is a genetic blood disorder that affects millions of people worldwide and is a major cause of illness and death in children, specifically in Sub-Saharan Africa. Previous studies have described changes in facial and craniofacial development among people with SCD but many of these changes require special assessment or medical imaging. We examined whether measurements obtained from standardized smartphone photographs could quantify facial differences associated with SCD. We collected 308 front-facing photos of children with SCD and matched healthy controls between 5 months and 19 years old. Using computer-based facial analysis, we measured a range of geometric and texture features and used interpretable machine-learning methods to determine which ones best distinguished children with SCD. The model identified a small set of geometric facial measurements that significantly differed between groups, with larger differences observed in several older age groups. The findings provide a basis for future longitudinal and externally validated studies of facial morphology as a potential digital biomarker of disease-associated change.

## Introduction

Sickle cell disease (SCD) is the most common inherited genetic disorder, affecting an estimated eight million people worldwide, with nearly half a million affected births each year ^1–3^. More than three-quarters of those living with SCD are of African and South Asian ancestry. In Sub-Saharan Africa, including the Democratic Republic of the Congo (DRC), limited access to newborn screening, confirmatory testing, and comprehensive care contributes to substantial childhood mortality and morbidity. These gaps are particularly consequential in settings with high burden of comorbidities and constrained health-system resources^4,5^.

SCD is a group of chronic and progressive inherited red blood disorders caused by various mutations in the beta-globin gene, resulting in abnormal red blood cells (RBCs) with sickle-like shape^6–9^. Sickle cell anemia, a severe manifestation of SCD, is caused when an individual inherits both copies of the mutation leading to hemoglobin SS genotype ^10–12^. Due to sickling and changes in cell formation, RBCs become less flexible and prone to rupture, leading to anemia. Recurrent hemolysis, anemia, Vaso-occlusion, and bone-marrow activity contribute to multisystem complications across life course.^8,11–14^. Craniofacial and maxillofacial changes have also been reported, including alterations associated with marrow growth and expansion^12,15–17^. However, subtle soft-tissue facial differences are difficult to quantify during routine clinical assessment, and their relationship to age and disease history remains incompletely characterized^18^. Leveraging novel technologies, quantitative facial phenotyping may therefore provide non-invasive method for studying disease-associated morphology and, alongside laboratory testing for diagnosis, may provide insights relevant to the identification of lifetime complications and long-term management of patients with SCD.

Diagnosis of SCD depends on hematological testing, including hemoglobin electrophoresis, molecular testing among others. Preventive care strategies, management and disease-modifying strategies can substantially improve outcomes when connected with longitudinal care and community-based support ^19–21^. Nevertheless, access to testing, specialist services, and repeated assessments remain uneven in many resource-limited settings^22^. Digital measures that can be obtained non-invasively and made widely available may eventually complement established clinical and laboratory assessment while helping prioritize future evaluation.

Applications of Artificial Intelligence (AI), such as machine learning (ML) and deep learning methods, have been increasingly introduced to aspects of SCD management and have shown the potential to enhance diagnosis, predict complications, and improve patient outcomes^23–27^. Furthermore, AI methods have been employed to interpret large, complex data, facilitating the detection of SCD through digital analysis^26^. Interpretable, facial-analysis methods have been used to quantify dysmorphic patterns associated with several genetic conditions including trisomy 21, 22q11.2 deletion syndrome, Williams-Beuren syndrome, Noonan syndrome and Cornelia de Lange syndrome ^28–32^. Prior work by our group has combined landmark-derived geometric measurements with local texture descriptors and has demonstrated the importance of evaluation these methods in populations that are underrepresented in algorithm development, including children in the DRC. The present study differs from prior syndrome-classification studies in two important respects: SCD is not primarily diagnosed by a congenital facial phenotype, and the facials changes of interest may reflect heterogenous, age-associated consequence of chronic disease. Accordingly, our emphasis is on quantitative phenotype characterization and preliminary internal discrimination rather than clinical diagnosis^29–33^.

Therefore, we sought to examine and quantify the facial feature characterization in an age- and sex-matched pediatric and young adult cohort from Kinshasa. We further evaluated the discriminative performance of an interpretable ML model and examined the cross-sectional age- and sex-associated variation in selected features. This study was designed as an exploratory feasibility analysis and the potential of the utility of a smartphone-based, explainable framework with potential relevance to resource-limited global health settings.

## Results

To identify differential facial features, we applied our ML analysis pipeline to 308 participants (154 electrophoresis-confirmed SCD and 154 age- and sex-matched controls). Of the 73 prespecified facial features **Fig.1**, 14 differed between SCD and control (q<0.05, two-sided Mann-Whitney U-test after Benjamini-Hochberg multiple testing correction for false discovery rate [FDR]). Of the 14 significant features presented in **Table 1**, eight were geometric measures, i.e., distances and angles between facial landmarks, and six landmark-centered texture descriptors. Seven of the geometric measures were lower in the SCD cohort compared to controls (q<0.05): the distance between medial and lateral canthi, distance from nasion to philtrum, distance from columella to lower lip, distance from medial canthi and columella, distance between nose alae, nose length (distance between the nasion and columella), and angle at nasal alae. Only the distance between the medial canthi was greater in individuals with SCD than in controls (q<0.001). The six significant texture descriptors features were concentrated around the nasal ale, nasal root, columella, philtrum and upper lip.

**Fig 1.**
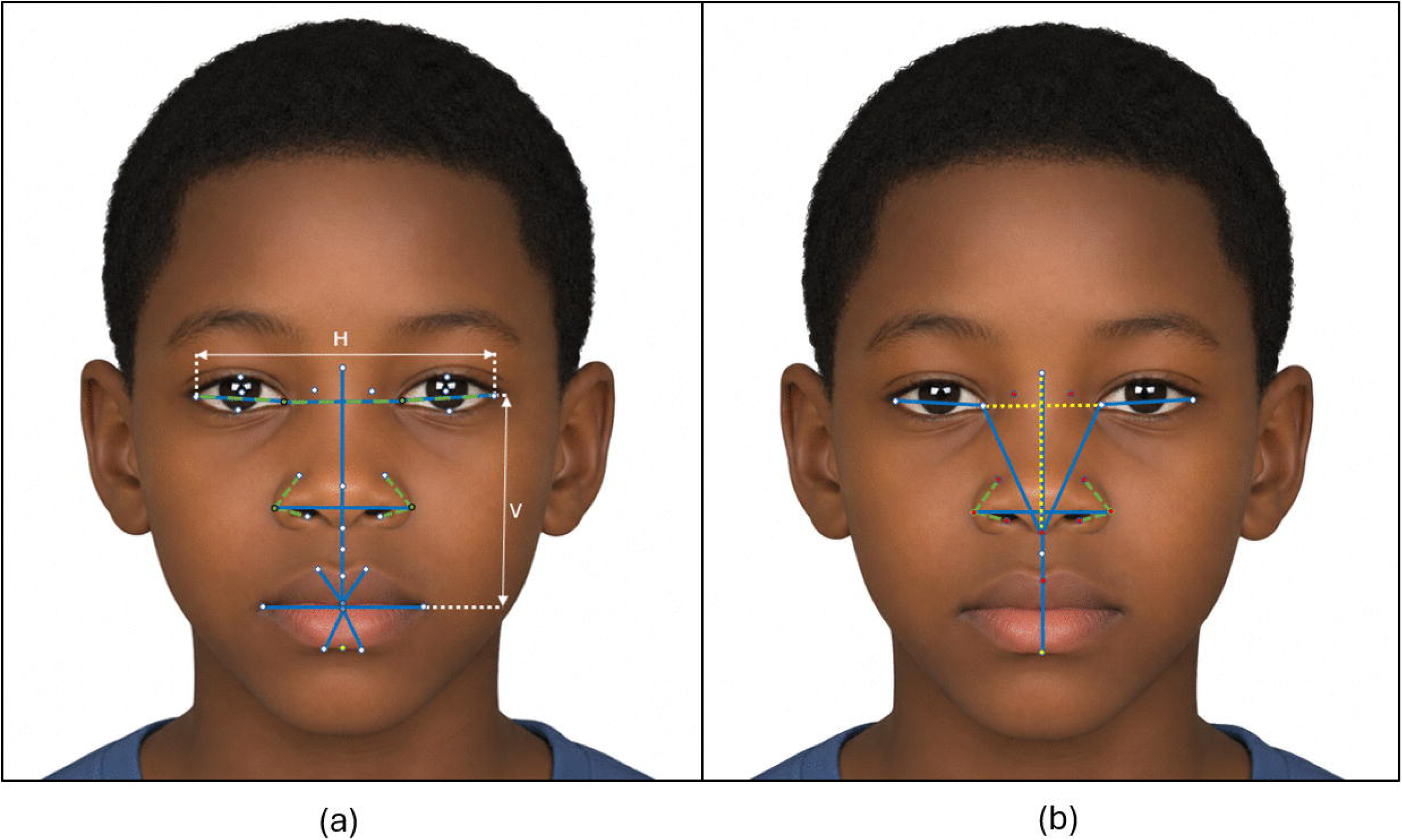
Illustration of landmarks and facial features. a) All facial landmarks and features used in the study. Blue solid lines represent the 11 distance features, part of the geometric features. Horizontal distances were normalized with respect to the distance between the lateral canthi (H), and vertical distances were normalized to the distance between the lateral canthi and the oral commissures (V). Dashed green lines represent the two angle features, part of the geometric features. The green dots represent the points at which the angles are calculated. Appearance features were calculated with different radii around each of the 33 landmarks represented as white and green dots. The yellow dot at the midpoint between two landmarks on the lower lip was included only for defining distance features. A list of all features and landmarks metrics can be found in **Data File S1. b**). **Discriminative features identified by the machine learning model.** Blue solid (distance) and green dashed (angles) lines represent the six discriminative geometric features selected by the ML model for SCD identification (see **Table 1**). Yellow dashed lines and red dots represent two additional geometric features and six appearance features that were significantly different between the SCD and control cohorts but were not selected by the ML model (see **Table 1**). Illustration does not depict or represent any real study participant.

**Table 1.**
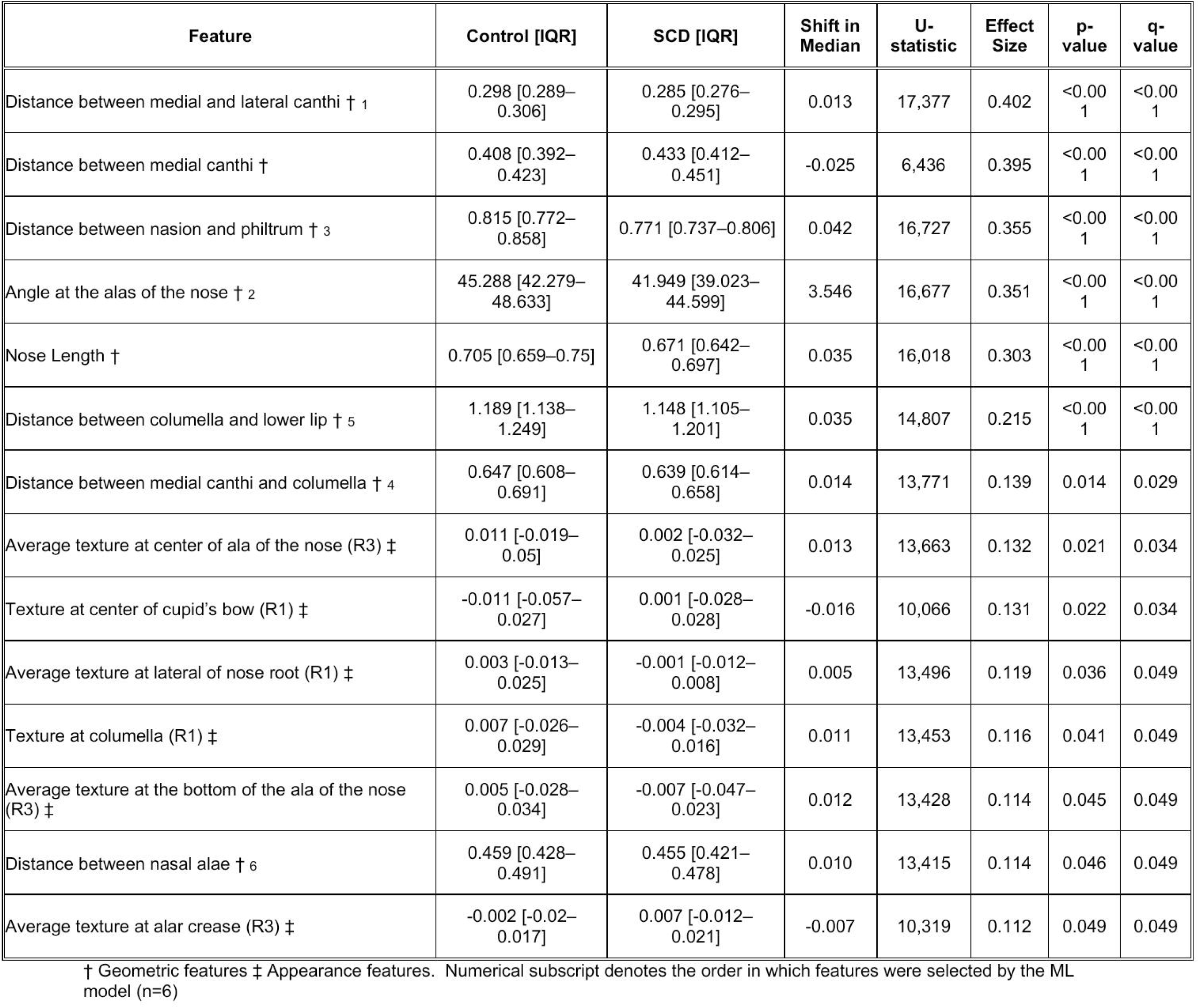
Feature comparisons retained in the internally validated model discriminate between normative controls and SCD patients. The list is arranged in descending order of the discriminatory power. R1=4 pixels, R2=8 pixels, and R3=12 pixels refer to the three radii used to calculate each appearance feature. Statistical comparisons between the normative and SCD cohorts were done using the two-sided Mann-Whitney U-Test, where q-values represent adjusted p-values using the Benjamini-Hochberg method for multiple testing correction FDR. IQR represents the interquartile range. Effect size represents the rank-based effect size 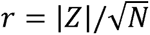. where Z is the standardized Mann-Whitney U statistic and N is the total sample size for the comparison.

Within the leave-one-out cross validation ML model, six features, all geometric, were frequently retained and achieved the highest discriminatory accuracy between SCD and control groups. These features were, in order of discriminatory power: the distance between medial and lateral canthi, angle at nasal ala, distance from nasion to philtrum, distance from medial canthi to the columella, distance from columella to the lower lip, and distance between nasal alae. The ML model achieved an SCD discrimination accuracy of 0.795 [95%CI:0.750,0.840] and an area under the receiver operating characteristic curve (AUROC) of 0.843 [95%CI:0.798,0.887] with sensitivity and specificity of 0.812[95%CI:0.750,0.871] and 0.779 [95%CI:0.707,0.838] respectively. The model yielded a PPV of 0.788, a NPV of 0.811, a F1-Score of 0.803, and a balanced accuracy of 0.799 at a classification threshold of 0.5. The corresponding confusion matrix, summarizing true-positive, true-negative, false-positive, and false-negative classifications is presented in **Fig S1.** Definitions and values for all geometric and appearance features evaluated in the analysis are provided in **Data File S1.**

Six texture descriptors differed at varying radii around the landmarks (R1=4 pixels, R2=8 pixels, and R3=12 pixels for facial images of standardized to 150×150 pixels), however, no appearance features were selected by the ML model in our discriminatory analysis (**Table 1**). **Fig. 2** illustrates the significant appearance features (q<0.05), which in general concentrated in the regions surrounding the philtrum, nasal alae, columella, upper lip and nose root. The texture figure was created by extracting grayscale patches centered on the selected landmark from all preprocessed facial images and averaging them pixel-wise within each group. For preprocessing details, see Material and Methods sections.

**Fig 2.**
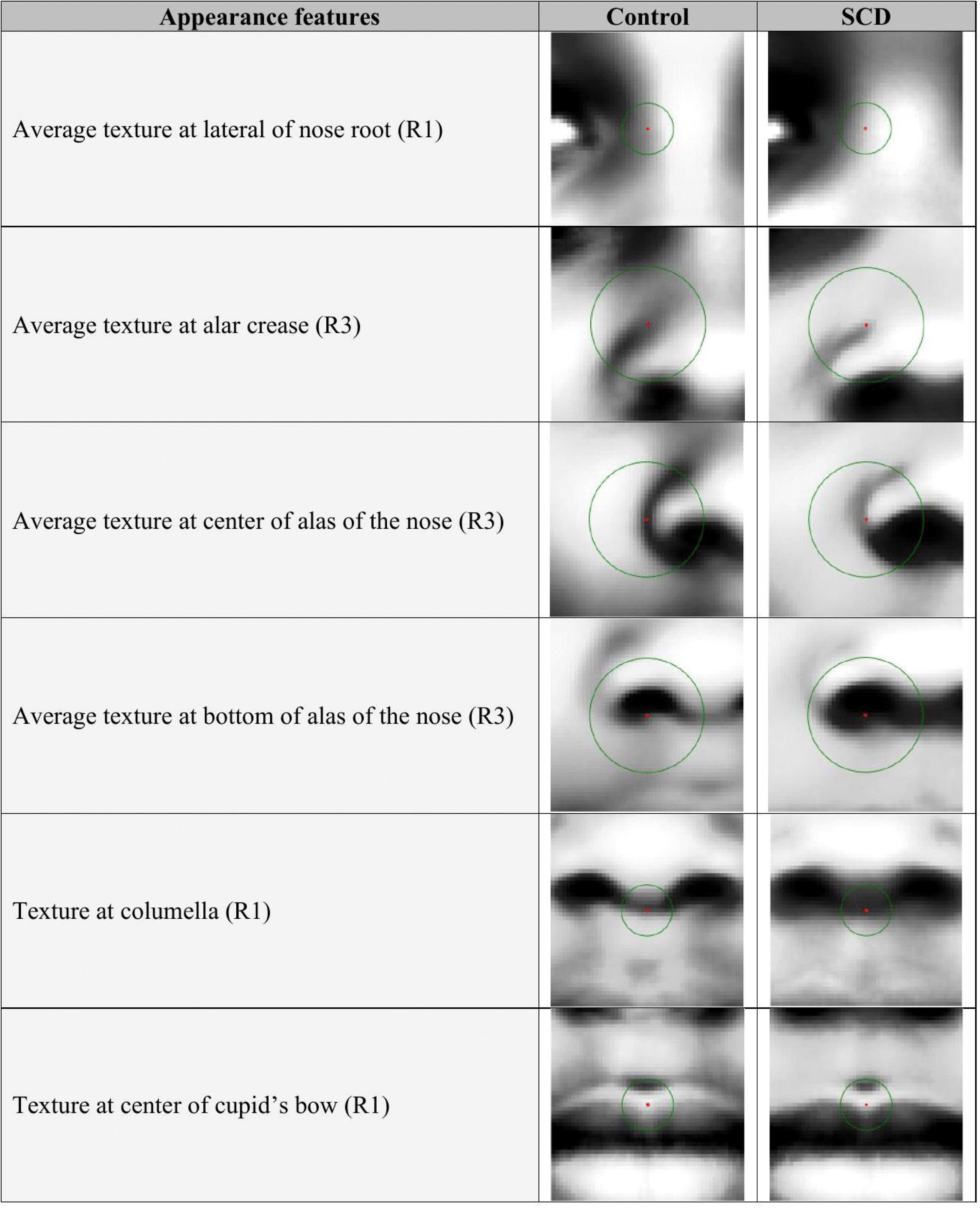
Discriminative appearance features. Cohort-level synthetic generated average and normalized. representations of the significant appearance features are presented for the control and SCD cohorts. R1=4 pixels, R2=8 pixels, and R3=12 pixels refer to the three radii used to calculate each appearance feature. Green circles represent the area around the landmark (in red) that is involved in the calculation of the appearance feature.

Cross-sectional age-stratified comparisons were conducted. Several SCD-control differences were smaller in younger groups (**Fig. 3)**. The intercanthal distance did not differ in the youngest age groups (1-2) but was reduced in SCD at age group 3 (q=0.034) and further accentuated at age groups 4 and 5 (q<0.001). Similarly, the angle at the alae of the nose exhibited significant changes in SCD compared to controls in age groups 2 through 5 (age group 2-4 q=0.007, age group 5: q<0.001). The distance between the nasion and philtrum was significantly reduced within the SCD cohort in age group 3 (q=0.003) and observed in age groups 4:5 (q<0.001, U-test) compared to controls. The distance between the columella and lower lip exhibited differences in age group 4 (q=0.031) and was accentuated in age group five (q<0.001, U-test). The medial canthi to columella only exhibited a pronounced difference in age group 5 (q<0.001), while nose alae distances did not significantly differ by age between our cohorts. The cross-sectional nature of these differences indicates age-associated differences and not within-person progression.

**Fig 3.**
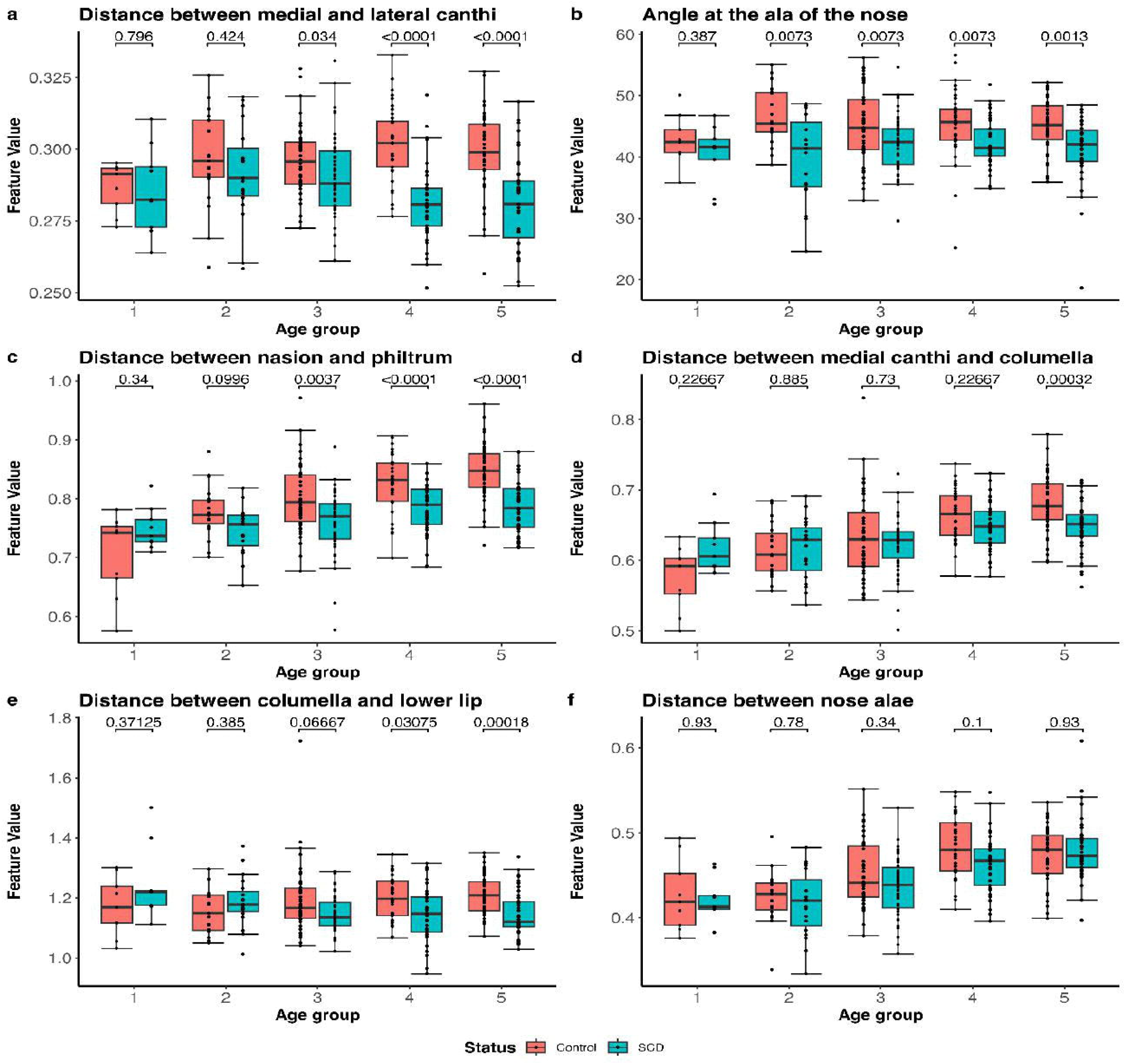
Age-stratified comparisons of discriminative geometric features. Boxplot representations of selected facial features are presented between individuals with SCD and controls. Each plot illustrates feature values across the five age groups (1–5). Features include the distances between the medial canthi and lateral canthi, medial canthi and columella, columella and lower lip, nasion and philtrum, nasal alae, as well as the angle at the alae of the nose. For each box, the median value is illustrated, the lower and upper edges of the box represent the 25^th^ and 75^th^ percentiles, and whiskers indicate the interquartile ranges. Data points beyond the whiskers indicate outliers. Statistical comparisons between control and SCD groups were done using the two-sided Mann-Whitney U-Test adjusted using the Benjamini-Hochberg method for multiple testing correction FDR. Q-values are presented for each compared group.

Although the prevalence of SCD does not appear to be sex related due to its autosomal-recessive transmission, progression and presentation may differ^33^. Of the six selected features, only two were significant by inspecting sex differences between females and males within disease status. The distance between the medial canthi and columella was smaller in SCD males than in SCD females (q=0.024), while the distance between nose alae was larger for males than for females in the control cohort (q=0.004). Further comparisons between SCD and control within sex identified statistical differences in all but two features among females: the distances between the medial canthi to the columella and between the nose alae (q=0.232 and q=0.083, respectively). Complete sex-stratified comparisons of the six geometric features by the ML model, including comparisons between females and males within each cohort and between SCD and control groups within sex, are presented in **Fig.S2**.

Together, the magnitudes of several between-group facial differences were greater in older age categories than in younger. To further illustrate these demographic variations in facial appearance by age and sex, a composite of discriminative facial features was composed to represent the differences in our comparison groups **Fig. 4**.

**Fig 4.**
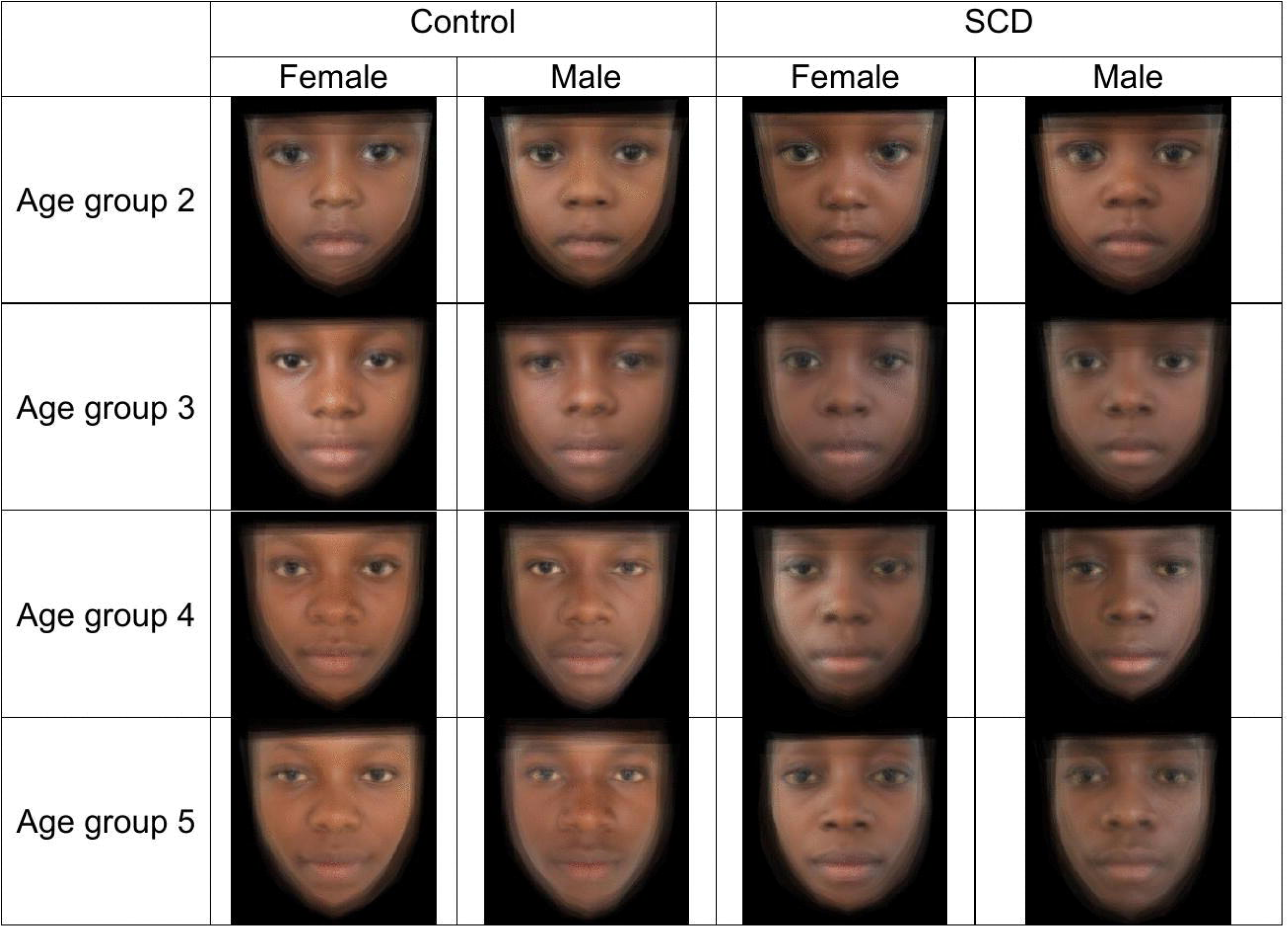
Composite average facial appearance and variability between controls and SCD samples by age and sex. Average faces were generated for each demographic group. Age group 1 (5 months to 3 years) was not included due to the insufficient sample size to create average facial representations. Cohort-level composite facial representations generated by spatially aligning and averaging images from multiple participants. The resulting composites do not depict any individual study participant.

## Discussion

In this exploratory study, quantitative facial phenotyping identified geometric and texture features associated with SCD in an age- and sex-matched cohort from Kinshasa. An interpretable linear model based on a small set of geometric measurements showed moderate internal discrimination, and several varying between-group differences. These findings support the feasibility of studying SCD-associated facial morphology in typically underrepresented environments^3,19,34,35^. Home to one of the largest populations of SCD outside of Nigeria and India, the DRC faces a multitude of challenges in the systematic screening and care of SCD patients^36^. The high burden of comorbidities, economic disparities, and lack of birth and death registries underscore the difficulties in the management and treatment of SCD in the DRC^4,19,36,37^. This setting provides an important context for developing research methods that rely on accessible acquisition tools. Adaptive and easily implemented technologies, like smartphone photography and ML capacity, may lower the burden of the collection of standardized morphometric data providing avenues for the development of digital technologies implementable in low resource settings.

In recent years, the implementation of facial analysis technologies in healthcare has increased. Its utilization in genetic conditions, such as Trisomy 21 and Noonan syndrome, has been extensively characterized ^28,30,31,34,35^. However, facial analysis has not been broadly studied in SCD, likely due to the subtlety of the facial patterns observed with the disease. Previous studies on SCD morphology have largely concentrated on hematological parameters and skeletal changes, particularly in the cranial and maxillofacial bones due to bone marrow expansion^38–42^. This study extends this literature by quantifying soft-tissue geometric and local texture features from standardized frontal photographs and by using an interpretable ML model to examine combined discriminatory information. Our selection of a support vector machine (SVM) with recursive feature elimination followed the same interpretable, geometric-feature-based framework validated in our team’s prior work across diverse and underrepresented populations, specifically including Trisomy 21 in the DRC^31^, demonstrating that this approach performs reliably well in small to medium cohorts without requiring deep-learning scale data. The contribution is therefore not the replacement of established testing nor as a standalone clinical tool, but the development of an interpretable and reproducible framework for studying facial morphology alongside standard clinical procedures in a population and setting underrepresented in digital-health research.

In this study, we identified 14 discriminative geometric and appearance features of the face that are associated with SCD. Our ML model based on facial analysis achieved an AUROC of 84.3% for the automated identification of children with SCD from non-invasive, widely available photography. The model’s AUROC and balanced sensitivity and specificity indicate that features contain meaningful discriminatory information for this cohort but does not support standalone clinical use.

Six geometric features were selected by the ML model as being the most discriminatory for SCD identification. The selected geometric features predominantly characterize the periocular, nasal and perioral regions of the face, suggesting that the SCD-associated phenotype reflects differences across multiple components of facial morphology rather than a single anatomical measure. Most notably, we identified reduced distances between the medial to lateral canthal distance, and lateral canthi, between the medial canthi and columella, and intermedial-canthus distance in SCD subjects compared to normative controls^17,38^. Pithon et al. found changes in the mandibular setback and inclination of the incisors of SCD patients^43^. These changes differed with age, with older individuals exhibiting more pronounced features, a trend also observed in our study. These patterns may reflect age-associated disease exposure, cohort differences, treatment history, selective survival or other unmeasured factors, however within-person morphological progression association cannot be elucidated within this study. Feature variations were most pronounced in older age groups within our post-hoc cross-sectional age analysis. These patterns are broadly consistent with prior reports of altered craniofacial and maxillofacial morphology in SCD, although direct comparison is limited by methodological and population differences ^17,38^.

This investigation also identified texture-based differences in regions around the nose and upper lip, however these texture differences were not selected by the final discriminatory model. Local binary-pattern features summarized image contrast and spatial variation however, more robust studies including standardized color calibration, repeated imaging and clinical covariates applied to facial texture analysis in SCD patients may reveal new insights into disease-associated physiological, pathological, and neurological changes. Variations in skin texture may reflect changes associated with vaso-occlusive crises, chronic anemia, or systemic inflammation, as these conditions alter vascular dynamics and tissue oxygenation^14,39,44,45^.

AI and ML approaches in SCD have increasingly expanded beyond automated disease detection toward the prediction and characterization of disease severity, complications, and clinical outcomes^40,46,47^. Early applications included automated classification of sickled erythrocytes from digital blood-smear images and image-based detection of sickle cell retinopathy, while other models have been developed to predict hospital readmission, acute care utilization, and the need for intervention in SCD-related complications ^48^. More recent studies have integrated clinical, laboratory, imaging, and electronic health record data to predict disease related events, complications and severity, further demonstrating the potential of data-driven methods to support individualized assessment of SCD ^24,49,50^. Explainable ML approaches may be particularly valuable in SCD, where the clinical phenotype is heterogeneous and evolves through the cumulative effects of disease associated factors. Within this emerging framework, our study extends explainable ML methods to quantitative facial phenotyping. To our knowledge, this is the first study to apply ML to the morphometric analysis of SCD and provides an avenue for investigating measurable phenotypic features of the disease.

This study is not without limitations. The composition of both study groups introduces additional uncertainty and may reflect clinical heterogeneity within the cohort while the single site nature limits the representativeness across regions and ancestry. The cross-sectional nature limits the ability to assess influences on facial morphology and growth. Clinical factors including hemoglobin genotype, anemia severity, transfusion history and other cumulative complications were not available to researchers. Furthermore, controls were considered non-SCD by local clinicians however did not undergo confirmatory electrophoresis. If trait status influences facial morphology, inclusion among controls, may reduce between group separation. More broadly, this study was conducted in a single-site cohort and relied on internal cross-validation without external validation, potentially limiting the assessment of generalizability to other populations and settings. The cross-sectional nature of the study may also reflect cohort and survival effects that may confound any assumptions of developmental progression^58^. The uniqueness of the DRC and ethnic diversity in the population in Kinshasa may also present a limitation for the broader implications of this investigation^51^. Nevertheless, by carefully matching the control and SCD cohort and using explainable ML methods appropriate for small datasets, we identified, for the first time, significant and distinct facial features of SCD and age-associated difference of the disease that were more pronounced in older age groups.

The integration of ML methods in SCD research offers promising opportunities. Future studies incorporating explainable ML methodologies for SCD should include prospective enrollment, repeated longitudinal imaging and detailed measures of disease complications and treatment. Incorporation of larger datasets may also permit comparison across demographically different groups and influence external validation. Within that framework interpretable facial features could be utilized as a component of a multimodal research tool incorporating laboratory and clinical information. Such approaches, successfully applied to other genetic conditions with facial dysmorphology^52–54^, could enable non-invasive monitoring of disease progression and early detection of complications, such as tissue remodeling seen in pathology of SCD. By adopting an open science framework, with publicly available data and methods, this work aims to support continued investigation and innovation in the field.

## Conclusion

In summary, interpretable ML-driven facial analysis technology that automatically extracts a suite of quantifiable morphometric biomarkers from smartphone photographs, revealed distinctive facial characteristics associated with SCD in an underrepresented cohort. Several geometric differences were pronounced in older age groups, although the cross-sectional design does not establish individual progression. To our knowledge, this is the first ML-based study of the facial morphology associated with SCD. These findings provide a foundation for longitudinal, multisite and clinically characterized studies evaluating facial morphology as a potential digital biomarker within a broader supportive assessment framework.

## Materials and Methods

### Ethics Statement

Ethical approvals for this study were obtained from the National Health Ethics Committee of Kinshasa, Democratic Republic of the Congo (IRB: #418). Prior to recruitment, DRC-based pediatricians and researchers visited the *Centre de Medecine Mixte et d’Anémie SS* (CMMASS) and held information sessions with nurses and administrators on the scope and goals of the study and subsequently acquired approval to conduct the study at this site. Informed consent was obtained from all participants or from a parent or guardian, with assent obtained from minors when appropriate. Consent procedures were conducted in the participant’s preferred language (French or Lingala). When literacy limitations prevented written documentation, verbal consent was obtained. Participants were explicitly informed about the collection and use of facial images for research purposes. Consent for participation did not automatically authorize public release of identifiable images, only data permitted by the approved consent and applicable privacy requirements will be shared. All participants were provided with a copy of their consent form.

### Study Design

From 2019 to 2023, participants were recruited in Kinshasa through convenience sampling. Participants with SCD were recruited from CMMASS, the primary specialized care clinic for patients with hematological conditions in Kinshasa, and were eligible if SCD had been confirmed by gel electrophoresis. Normative age- and sex-matched controls were collected from the local community and were considered not to have SCD based on assessment by our local clinicians. Each participant was matched 1:1 to a control by sex and age. Additional matching was not performed. The research protocol and aims were explained in detail in the local language to parents and participants prior to obtaining assent or consent for participation. Following informed consent, front-facing facial photographs and demographic information of the participants were captured using Facial photographic capture smartphone app used in conjunction with the mGene software for facial analysis^34^. In total, 154 children diagnosed with SCD and 154 age- and sex-matched controls aged 5 months to 19 years were included in this study. There were 154 males (50%, mean age 11.31 ± 6.35 years) and 154 females (50%, mean age 12.51 ± 6.11). All participants were recruited in Kinshasa and were described as of African descent. The hemoglobin type of the confirmed SCD cases was unknown.

### Image Acquisition

Front-facing photographs were acquired in a standardized format using mGene image software with iOS-supported mobile devices by trained research personnel. Participants were positioned seated or standing with capture areas (face) approximately 10 to 18 inches distance from the camera with any items obscuring or obstructing the face removed. Images were subsequently captured with the camera lens at eye level and centered when a neutral expression with eyes open and mouth closed was present. Whenever feasible, photographs were acquired under diffuse ambient lighting while avoiding direct sunlight, strong shadows, or backlighting. Research staff visually reviewed each image immediately after acquisition. Images were repeated if there was excessive head rotation or tilt, facial occlusion, motion blur, poor focus, inadequate illumination, or incomplete visualization of facial boundaries. Prior to any selection into image analysis, secondary quality control checks were performed by the US-based study team. The in-country research team was contacted to recapture photos if any of the prespecified criteria were not met; otherwise, photos were excluded.

### Facial Analysis

Digital facial analysis technology was used to evaluate the front-facing photos of individuals with SCD and their age- and sex-matched controls using methods developed by our team and previously described^28–31^. For preprocessing, 33 facial landmarks were manually annotated on each image. The landmarks were used to perform a rigid landmark-based registration to a reference face using a least-squares point-set alignment, placing all faces into a common coordinate system. Registered images were sampled to a fixed image size, converted to a grayscale, and cropped to a standardized 150×150-pixel resolution prior to feature extraction^35^. For feature calculation, our technology identified and quantified 13 geometric features, including 11 distance features and 2 angle features, from 33 anatomical facial landmarks (e.g., lateral canthi, oral commissures, etc. shown in **Fig. 1**). The distance features were normalized to the size of the face, e.g., horizontal distances were normalized with respect to the distance between the lateral canthi, and vertical distances were normalized to the distance between the lateral canthi and the oral commissures. In addition, the technology quantified the radiomic-style features around each of the 33 facial landmarks using a multi-resolution texture descriptor based on local binary patterns^30,35,55,56^, which are sensitive to lines, shadows, and local contrast. Among the 33 landmarks, 26 (13 pairs) are symmetrically placed on the left and right sides of the face, such as the lateral canthi. For these pairs, appearance features were calculated as the average at the left and right landmarks. To capture multi-scale patterns, appearance features were calculated with three radii (R1=4 pixels, R2=8 pixels, and R3=12 pixels), yielding a total of 60 appearance features (20 for each radius [13 pairs and 7 along the midline of the face]). **Fig. 1** illustrates the facial landmarks and the features defined from them. Together, these features quantify relative periocular, nasal, and perioral geometry and localized facial appearance, anatomical regions in which craniofacial and orofacial differences have previously been described in facial morphology of genetic conditions^25,30,34,35,38,41,52,57^. Area-based measures and explicit symmetry indices were not calculated in the present analysis. No feature-value imputation was performed. Images that failed landmark detection were excluded before analysis.

Initial feature filtering was performed using the non-parametric two-sided Mann-Whitney U-test using Benjamini-Hochberg correction for multiple testing FDR ^58^ to compare feature values between controls and patients with SCD. Features with q>0.05 were excluded. From the remaining features, the most discriminative were selected using recursive feature elimination^59^ and a ML model adequate for a small to medium dataset, i.e., a support vector machine model with a linear kernel^60^. To select the optimal number of features for SCD identification and avoid overfitting, the number of selected features was progressively increased until the AUROC converged^61^. The ML model was trained on the selected features to generate a classification score for the discrimination between participants with SCD and controls based on their facial presentation. Model training and validation were performed using leave-one-out cross-validation. Within each leave-one-out iteration, statistical feature screening and recursive feature elimination were performed using only the training participants; the held-out participant was used only to generate the corresponding out-of-fold prediction. Model performance was not interpreted as external or prospective validation. We report AUROC, accuracy, sensitivity, specificity, positive predictive value, negative predictive value, F1-score, balanced accuracy and the confusion matrix.

### Cross-Sectional Analysis

Age- and sex-stratified analysis was conducted with two-sided Mann-Whitney U tests with Benjamini-Hochberg multiple testing correction for FDR to examine whether SCD-associated feature differences varied across age categories. Participants were grouped to represent developmental phases of human growth, ensuring alignment with the physiological and morphological changes that occur at each stage. These age categories capture key transitions, such as the rapid growth during infancy and toddlerhood (age group 1: 5 months–3 years, n=18), establishment of structural features in preschool age (age group 2: 3–6 years, n=38), and the rapid changes in craniofacial morphology during school age (age group 3: 6–13 years, n=92) and adolescence (age group 4: 13–18 years, n=74). The inclusion of young adults (age group 5: >18 years, n=86) allowed for the examination of fully developed craniofacial features.

## Data Availability

All data produced in the present study are available upon reasonable request to the authors

## List of Supplementary Materials

**Fig. S1**. Confusion matrix of SCD classification using the leave-one-out cross-validation model.

**Fig. S2**. Sex-stratified comparison of geometric facial features selected by the machine-learning model. Comparisons are presented between females and males within the SCD and control cohorts and between SCD and control groups stratified by sex.

**Data File S1**. Facial morphometric features evaluated in the study. File contains definitions and values.

## Acknowledgments

We would like to thank all the CMMASS patients and their families for participating in this study. We are grateful to the clinical staff who facilitated the completion of this study. Resources utilized in this study were developed in part, with the support of NIH award R33HD102988.

## Contributions

Conceptualization: DS, XL, DMN, EV, DTK, MGL.

Data Curation: DS, XL, KM, GK, MSB, EL, LT

Methodology: DS, XL, MSB, DMN, EV, MGL

Formal Analysis: DS, XL, FM, MGL

Visualization: DS, XL, FM, MGL

Writing-Original Draft: DS, XL, MGL

Writing-Review & Editing: All Supervision: DTK, MGL

Corresponding Authors: MGL

**Correspondence** and request for materials should be addressed to Marius George Linguraru

## Data Availability

The raw facial morphometric data generated by this study are available in **Data File S1**. Source data are provided in our GitHub repository, as detailed below.

## Image Creation

Figures 1 and 2 are synthetic graphical representations created for illustration and do not depict real study participants. Figure 4 consists of cohort-level composite facial representations generated by spatially aligning and averaging images from multiple participants and does not depict any individual study participant.

## Code Availability

In alignment with open science principles, the source code used in this study is publicly available to facilitate transparency, reproducibility, and support ancillary research initiatives. The code was written in Python and is available on GitHub at https://github.com/Pediatric-Accelerated-Intelligence-Lab/mGene-ml. The repository provides a feature analysis pipeline for processing images with labeled landmarks. The output is an Excel file that summarizes feature statistics, cross-validation performance, and the selected features. Additionally, the codebase includes modules for appearance feature visualization (creation of **Fig. 2**) and average face generation (creation of **Fig. 4**).

To demonstrate the capabilities of the software, the repository provides public data consisting of facial images of 20 SCD cases used in this study, all of whom have consented to the public release of their data. A comprehensive README.md file is included in the repository, detailing system requirements, installation guide, functional descriptions, demo data usage, and expected outputs.

---

**Figure S1: Confusion matrix of SCD classification using the leave-one-out cross-validation model.** The confusion matrix summarizes the classification of individuals with SCD and age- and sex-matched controls based on the six geometric facial features selected by the machine-learning model.

**Figure S2: Sex stratification and comparisons of the selected features by the machine learning model for sickle cell disease identification.** Sex stratified analysis was conducted for each of the selected features in our cross-validation analysis (control males, n=77; control females, n=77; SCD males, n=77; and SCD females, n=77). **A) Within control and SCD group comparisons. B) Between control and SCD group comparisons.** For each box, the median value is illustrated, the lower and upper edges of the box represent the 25^th^ and 75^th^ percentiles, and whiskers indicate the interquartile ranges. Data points beyond the whiskers indicate outliers. Statistical comparisons were done using the two-sided Mann-Whitney U-Test, and q-values show adjusted p-values using the Benjamini-Hochberg method for multiple testing correction FDR.

