## Supplementary figures and images for "Quantitative Facial Phenotyping and Age-Associated Differences in Sickle Cell Disease Using Interpretable Machine Learning"

### Supplemental Figure 1

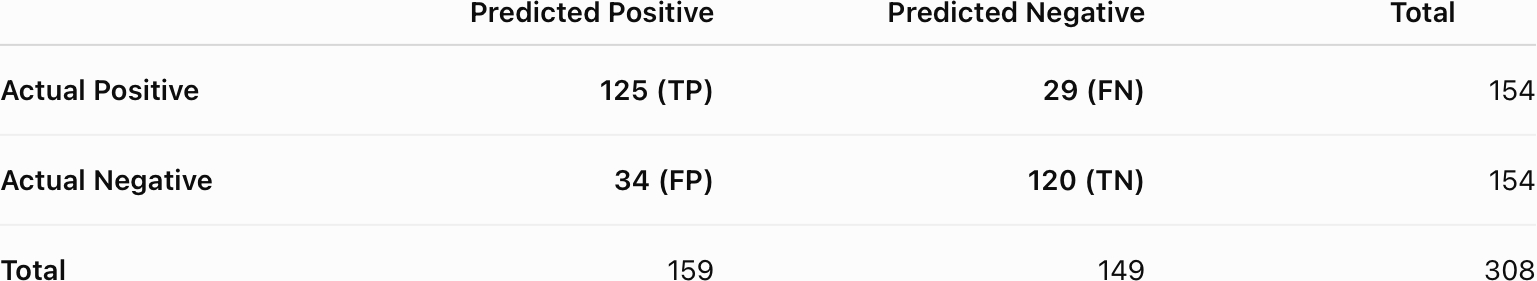

### Supplemental Figure 2

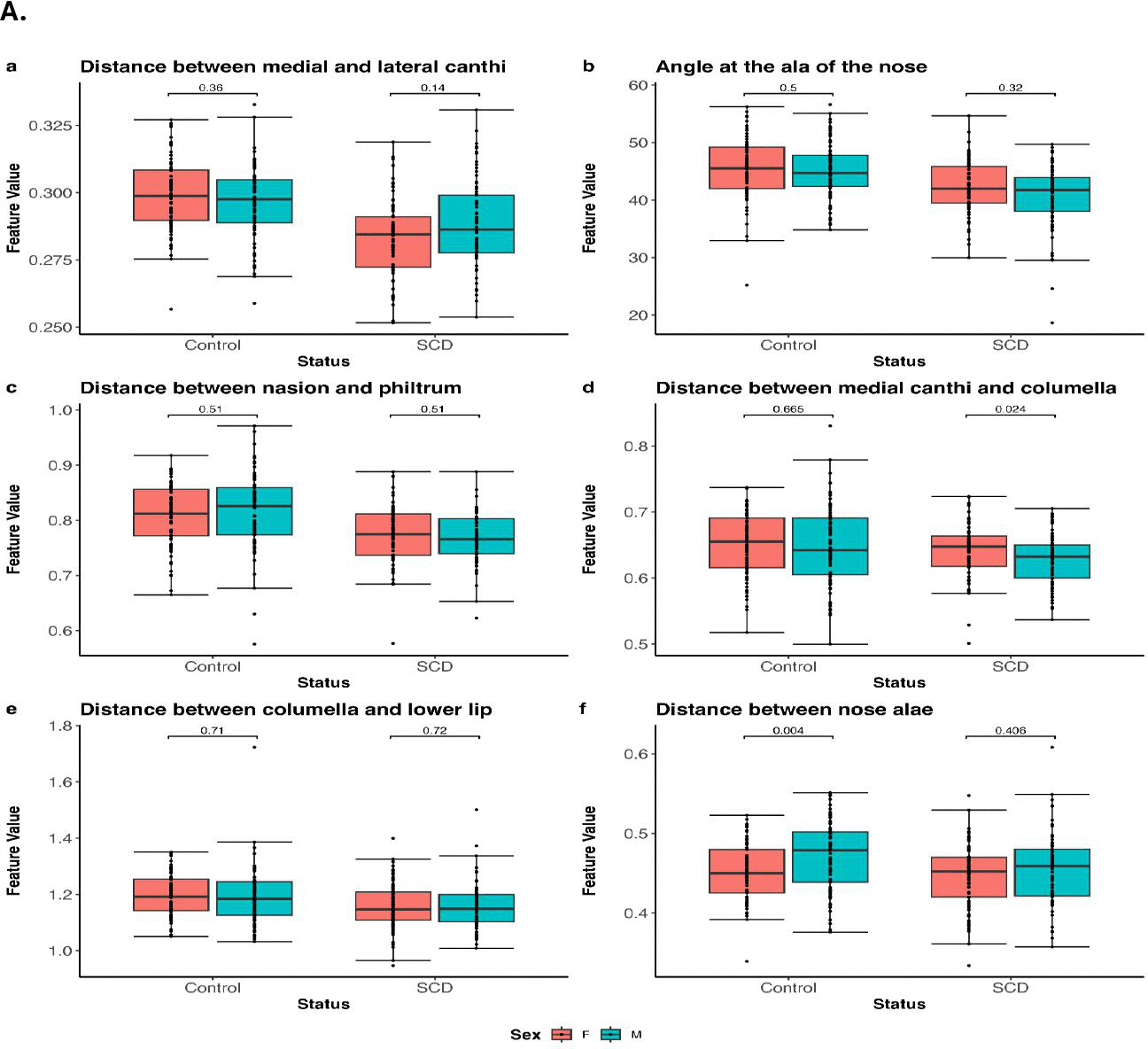

### Supplemental Figure 3

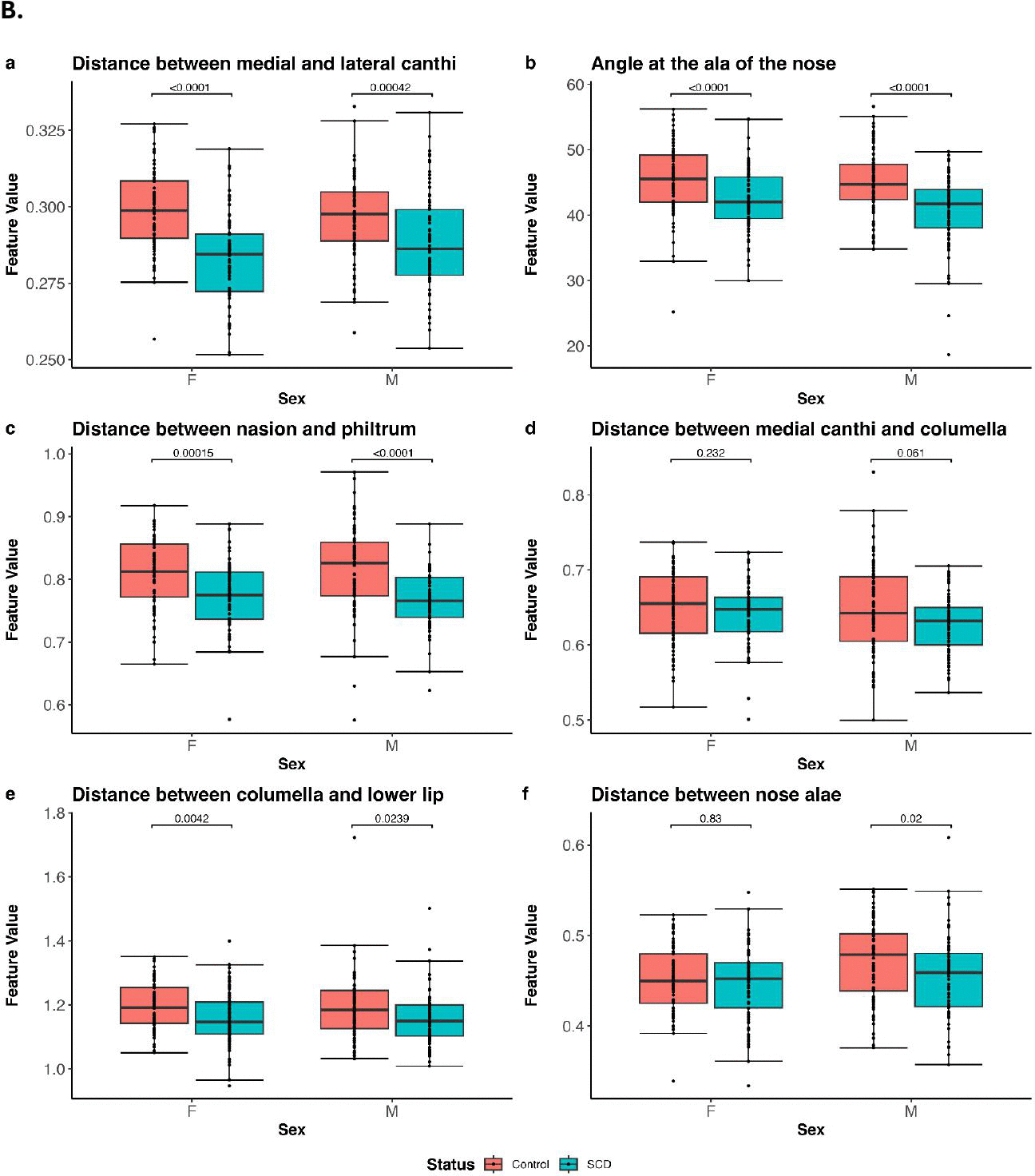
